# Personal network inference identifies children at risk of recurrent wheezing and asthma

**DOI:** 10.1101/2025.09.26.25336702

**Authors:** LA Coleman, SK Khoo, JA Bizzintino, K Franks, F Prastanti, M Borland, PN Le Souëf, DG Hancock, YV Karpievitch, A Bosco, IA Laing

## Abstract

**Background:** Wheezing and asthma exacerbations are leading causes of pediatric hospital admissions. Predicting which children will experience persistent exacerbations remains challenging. Prior research has identified immune endotypes in the nasal epithelium of children with acute asthma and wheezing, characterized by varying balances of interferons and inflammatory markers. Notably, children exhibiting low interferon responses coupled with high inflammation are at an increased risk for recurrent respiratory exacerbations.

**Objective:** This study aims to determine if blood-based gene network biomarkers can detect immune endotypes in children presenting with acute wheeze and asthma, potentially serving as predictive tools for future exacerbations.

**Methods:** We conducted gene expression analysis using microarrays of peripheral blood mononuclear cell (PBMC) samples from pediatric patients who presented to hospital for acute wheeze and asthma. Personal network inference was employed to discern gene expression patterns, facilitating the classification of patients into distinct immune endotypes.

**Results:** Three immune endotypes were identified. One endotype, characterized by low interferon responses and elevated expression of both innate and adaptive immune pathways, was significantly associated with an increased risk of subsequent hospital respiratory presentations and a persistent pattern of respiratory exacerbations over time.

**Conclusion:** PBMC-based personal gene network biomarkers can effectively identify immune endotypes correlating with clinical outcomes in pediatric asthma. The high-risk endotype represents a potential treatable trait in acute wheezing episodes. Therapeutic strategies aimed at enhancing interferon responses and/or reducing inflammation may benefit this subgroup.

**Clinical Implications:** We have identified a potential treatable trait of paediatric asthma and wheezing. Classifying children based on their immune profiles may enable tailored management strategies aligned with their future exacerbation risk.

**Capsule Summary:** Personal gene network biomarkers effectively identifies immune endotypes correlating with clinical outcomes in pediatric asthma. The high-risk endotype, marked by low interferon responses and high innate and adaptive inflammation represents a potential treatable trait in acute wheezing episodes.

## Introduction

Wheezing remains one of the most common reasons children present to hospital emergency departments, usually following a respiratory viral infection. Children under the age of fifteen accounted for 43% of hospitalisations for asthma in Australia in 2020-2021 (1) yet this is likely underestimated given children under 5 are less likely to be diagnosed with asthma (2). Asthma ranks among the top contributors to disease burden, defined in disability-adjusted life years for infants and young children (0-5 years), school-aged children (5-14 years) and young adults (15-24 years) (3). Paediatric asthma also imposes significant economic burdens, with children and adolescents under 19 years old accounting for nearly half (48%) of asthma-related public hospital emergency department costs and over a third of the total costs for public hospital admissions for asthma in Australia in 2019-2020 (4).

An area of intense research for improving the treatment and management of childhood wheezing and asthma exacerbation is identifying children at risk of developing persistent asthma, as opposed to children who will experience one or more wheezing episodes caused by respiratory infections that resolve with increasing age (2, 5, 6). The ability to distinguish between transient early wheeze and persistent wheeze and asthma is important clinically, as asthmatic children with a history of multiple severe exacerbations have been shown to have lung function deficits in later childhood and adulthood, making appropriate ongoing treatment a priority for this group (7). There is also a risk of inappropriately treating a sub-set of young children with corticosteroids, which can have systemic side effects such as reduction of growth (8). The precise mechanisms underlying persistence and remission are not well understood. One study of factors associated with increased risk of recurrent wheeze and asthma suggested potential roles for the respiratory microbiome, atopy, Th2 inflammation and RV-C; however, the potential mechanisms of these factor are not well understood (9).

We have previously employed personal network inference to unveil molecular subphenotypes underlying virus-induced acute lower respiratory illness in children (particularly acute wheezing and asthma) in nasal epithelial samples (10). We found that children with acute lower respiratory illness could be divided into endotypes based on their patterns of gene network expression. Those with networks showing deficient interferon response and dominated by inflammatory patterns were at greater risk of subsequent respiratory hospital exacerbations.

Now we aimed to examine systemic responses to viral infection using gene expression microarrays of peripheral blood mononuclear cell (PBMC) samples from a subset of children in the same cohort, investigating the presence of distinct immune endotypes. We hypothesised that children with acute lower respiratory illness could be categorised into specific endotypes based on their PBMC gene expression patterns, including profiles characterised into interferon-high/inflammation-low and interferon-low/inflammation-high as previously observed.

## Methods

### Participants and cell samples

Children aged 0-18 years were recruited into the Mechanisms of Acute Viral Respiratory Infection in Children (MAVRIC) cohort upon presentation to the emergency department of Princess Margaret Hospital (PMH) and later Perth Childrens Hospital (PCH) with an acute lower respiratory tract illness (ALRI), particularly acute wheezing and asthma (the samples used in this study were collected between 2009-2012). This study was granted ethics approval from PMH/PCH (Ethics #1761EP/RGS2369) and informed consent was obtained from at least one parent/guardian prior to recruitment.

At recruitment, children completed a skin prick test, and blood samples and nasal samples were collected. Asthma exacerbation severity score, atopy, total and specific IgE were assessed as described previously (11). The presence of common respiratory viruses (RV, RSV, adenovirus, influenza A and B, parainfluenza 1–4b and metapneumovirus) and specific pathogenic bacteria were assessed using a tandem multiplex RT-PCR assay (12). RV species were identified by genotyping a 270-bp variable sequence in the 5’ non-coding region of the RV genome, and RV species and genotypes were assigned as previously described (13).

### Recurrence data collection and phenotyping

Information on history of public hospital presentations and admissions from birth until study completion for a respiratory diagnosis was obtained from iSoft Clinical Manager (iCM), which is a public hospital database of hospital visits and test results. These data were used to characterise the longitudinal hospital respiratory exacerbation phenotype of participants as few/no exacerbations, multiple exacerbations or persistent exacerbations (14). Few exacerbations was defined as three or fewer hospital presentations during the observation period, with one or no presentations in the last 3.5 years and no diagnoses of acute asthma over the age of 5.5 years; multiple exacerbations was defined as four or more presentations over the observation period, with one or no presentations in the last 3.5 years and no diagnoses of acute asthma over the age of 5.5 years; and persistent exacerbations was defined as six or more presentations over the observation period, with multiple presentations in the last 3.5 years and at least one presentation diagnosed as acute asthma over the age of 5.5 years. Participants who could not be clearly classified into one of these severity groups due to possibly incomplete follow-up or unusual exacerbation patterns were termed “Could not be classified”.

### Transcriptomic analysis

Peripheral blood specimens were collected for extraction of PBMC RNA. In a randomly selected subset of acute and convalescent samples, PBMC RNA was extracted using TRIzol (Invitrogen, Carlsbad, CA, USA) and the RNeasy kit (Qiagen, Hilden, Germany). The RNA was processed and hybridised to genome-wide gene expression micro-arrays (Human Gene ST2.1; Affymetrix, Santa Clara, CA, USA) (available from Gene Expression Omnibus GSE298367).

Microarray data on PBMC samples were analysed in the open-source statistical software R (www.r-project.org/ accessed on 28 July 2020). The microarray data were pre-processed employing the RMA algorithm, using custom mapping of microarray probe-sets to the genome (*hugene21sthsentrezgcdf* Version 19) (15). The quality of the microarray data was assessed using the R package *arrayQualityMetrics*, and seven low quality samples were removed from the analysis. The top 20% most variable genes by standard deviation were selected for further analysis. These highly variable genes were clustered using <u>F</u>uzzy clustering by <u>L</u>ocal <u>A</u>pproximation of <u>ME</u>mbership (FLAME) to identify gene modules (16). Functional pathways of the identified modules were examined using InnateDB (17) and DAVID (18) pathway over-representation analyses and examining the top pathways. Gene modules were also visualised using STRING protein interaction networks (19). Personalised networks were constructed for each individual subject including control and convalescence samples by applying Linear Interpolation to Obtain Network Estimates for Single Samples (*LIONESS*) using the *lionessR* package (20). The network edges were then filtered for stability using an N-1 perturbation method, where an additional sample was removed, and *LIONESS* repeated. The log fold change between the N-1 network compared with the N network was then calculated and used to assess edge stability. The top one percent of stable network edges by edge weight were plotted using the software Gephi (21).

The top one percent of network edges for each individual acute case were then classified based on FLAME module membership. The module proportions were standardised and investigated for the presence of endotypes using Latent Profile Analysis (LPA) (22). For the *mclust* implementation of LPA, the best model solution is the one that maximises the Bayesian Information Criterion (BIC).

### Statistics

Continuous clinical characteristics were assessed for statistical normalcy using Shapiro-Wilk tests, then compared using parametric or non-parametric ANOVA and post-hoc *t*-tests or Wilcoxon tests (with Benjamini-Hochberg multiple test correction) as appropriate, whereas categorical variables were compared using Fisher’s exact tests with post-hoc paired Fisher’s exact tests (with Benjamini-Hochberg multiple test correction). Survival analysis of time until next respiratory hospital presentation or admission was conducted using the *survival* (23) and *survminer* (24) R packages.

## Results

### Cohort characteristics

MAVRIC participants included in this analysis (N=87) had a median age of 2.36 years (range 0.08-3.99 years) and 68% were male. The proportion of children who had corticosteroid administered in the 24 hours prior to blood collection was 70%. The proportion of children given a discharge diagnosis of acute asthma, bronchiolitis or wheeze was 92% (other diagnoses included pneumonia, upper respiratory tract infection and lower respiratory tract infection), and children often had multiple diagnoses. The proportion of children with a common respiratory virus detected at recruitment was 93%; 22% had RV-A detected, 55% had RV-C, and 10% had RSV. The proportion of children with atopy was 76%, with 47% having aero-allergen-specific atopy. The proportion of children with a subsequent hospital respiratory exacerbation was 75%, with a median time to either re-presentation or end of observation of 203 days, and median total follow-up duration of 8.28 years (Table 1).

**Table 1:** Cohort characteristics. Clinical variables for case children are summarised as number and percentage or median and range as appropriate. Corticosteroid use was defined as systemic corticosteroid administration in the 24 hours prior to blood collection. Time to re-presentation does not include children who did not have a subsequent exacerbation, whereas duration of follow-up includes all children.

|  |  |
| --- | --- |
| **N** | 87 |
| **Age at recruitment in years: median (range)** | 2.36 (0.08-3.99) |
| **Male sex: N (%)** | 59 (68) |
| **Time in days between symptom onset and presentation: median (range)** | 2 (0-43) |
| **Time in hours between presentation and blood collection: median (range)** | 15.37 (0.22-47.67) |
| **Corticosteroid administered in previous 24 hours: n/N (%)** | 59/84 (70) |
| **Exacerbation severity Z score at recruitment: median (range)** | 0.32 (-2.23-2.02) |
| **Hours to discharge: median (range)** | 24.9 (1.3-242.1) |
| **Diagnosed with acute asthma, bronchiolitis or wheeze: N (%)** | 80 (92) |
| **Season at recruitment: N (%)** | Spring 17 (19)<br>Summer 2 (2)<br>Autumn 24 (28)<br>Winter 44 (51) |
| **Any common respiratory virus positive: N (%)** | 81 (93) |
| **RV-A positive: N (%)** | 19 (22) |
| **RV-B positive: N (%)** | 0 (0) |
| **RV-C positive: N (%)** | 48 (55) |
| **RSV positive: N (%)** | 9 (10) |
| **Atopy: N (%)** | 66 (76) |
| **Aero-allergen-specific atopy: N (%)** | 41 (47) |
| **Number of children with a subsequent hospital exacerbation: N (%)** | 65 (75) |
| **Time in days to re-presentation: median (range)** | 96 (1-2998) |
| **Total duration of follow-up in years: median (range)** | 8.28 (4.53-13.24) |

### Personal network construction, module quantification within networks and cluster detection

Using our personalised network approach (see Supplementary Materials), we identified that a variable volume, variable shape, equal orientation (VVE) model with three components - hereafter referred to as Clusters A, B and C - optimised the Bayseian Information Criterion.

### Differences in clinical characteristics between clusters

We next aimed to determine whether the three clusters of children defined from personal network inference exhibited distinct clinical characteristics. Comparative analysis revealed significant differences among the clusters in discharge diagnosis, virus detection, RV detection, RV-C detection and longitudinal hospital respiratory exacerbation phenotypes (Table 2). The clusters did not differ in age at recruitment, sex, time between symptom onset and presentation, time between presentation and blood collection, corticosteroid administration in the previous 24 hours to recruitment, exacerbation severity Z score at recruitment, time to discharge, season at recruitment, RV-A detection, RSV detection, atopy, or aero-allergen-specific atopy (Table 2).

**Table 2:** Cluster demographics. Case children were divided into three clusters (A, B and C) using the personalised networks method. Clinical variables are summairsed as number and percentage or medina and range as appropriate, and were compared using non-paramteric ANOVA or Fisher’s exact tests as appropriate, with post-hoc non-paramteric *t* tests or paired Fisher’s exact tests with Benjamini-Hochberg multiple test correction as needed.

|  | Cluster A | Cluster B | Cluster C | Overall *p* value |
| --- | --- | --- | --- | --- |
| **N** | 30 | 23 | 34 |  |
| **Age at recruitment in years: median (range)** | 2.74 (0.08-3.99) | 2.26 (0.15-3.96) | 2.13 (0.08-3.89) | 0.61 |
| **Male sex: N (%)** | 22 (73) | 16 (70) | 21 (62) | 0.60 |
| **Time in days between symptom onset and presentation: median (range)** | 2 (0-10) | 3 (0-43) | 2 (0-7) | 0.46 |
| **Time in hours between presentation and blood collection: median (range)** | 12.23 (0.25-43.67) | 19.65 (0.97-37.12) | 12.77 (0.22-47.67) | 0.47 |
| **Corticosteroid administered in previous 24 hours: n/N (%)** | 21 (70) | 14/21 (67) | 24/33 (73) | 0.95 |
| **Exacerbation severity Z score at recruitment: median (range)** | 0.743 (-1.801-2.015) | 0.320 (-1.210-1.591) | 0.743 (-0.953-2.015) | 0.38 |
| **Hours to discharge: median (range)** | 29.6 (1.3-138.6) | 23.1 (2.3-99.6) | 23.5 (4.3-242.1) | 0.68 |
| **Diagnosed with acute asthma, bronchiolitis or wheeze: n (%)** | 29 (97) | 18 (78) | 33 (97) | 0.03* |
| **Season at recruitment: N (%)** | Spring 6 (20)<br>Summer 1 (3)<br>Autumn 10 (33)<br>Winter 13 (44) | Spring 2 (9)<br>Summer 0 (0)<br>Autumn 6 (26)<br>Winter 15 (65) | Spring 9 (26)<br>Summer 1 (3)<br>Autumn 8 (24)<br>Winter 16 (47) | 0.54 |
| **Any common respiratory virus positive: N (%)** | 25 (83) | 22 (96) | 34 (100) | 0.01* |
| **RV positive: N (%)** | 19 (63) | 17 (74) | 31 (91)^ | 0.02* |
| **RV-A positive: N (%)** | 8 (26) | 4 (17) | 7 (21) | 0.80 |
| **RV-C positive: N (%)** | 11 (37) | 13 (57) | 24 (70)^ | 0.03* |
| **RSV positive: N (%)** | 2 (7) | 4 (17) | 3 (9) | 0.45 |
| **Atopy: N (%)** | 22 (73) | 18 (78) | 26 (76) | 0.90 |
| **Aero-allergen-specific atopy: N (%)** | 15 (50) | 10 (43) | 16 (47) | 0.96 |
| **Longitudinal hospital respiratory exacerbation phenotype: N (%)** | Few 16 (53)<br>Multiple 10 (33)<br>Persistent 2 (7)<br>CNBC 2 (7) | Few 6 (26)<br>Multiple 11 (48)<br>Persistent 4 (17)<br>CNBC 2 (9) | Few 6 (18)<br>Multiple 17 (50)<br>Persistent 10 (29)<br>CNBC 1 (3) | 0.03* |

\* = $p < 0.05$ in overall comparison, ^ = $p < 0.05$ compared with Cluster A in post-hoc Fisher's exact test with Benjamini-Hochberg multiple test correction, CNBC = could not be classified

Cluster B had a lower prevalence of children diagnosed with acute asthma, bronchiolitis or wheeze compared with Clusters A and C (Cluster A: 97%, Cluster B: 78%, Cluster C: 97%); however, this difference was not significant in post-hoc tests after adjusting for multiple comparisons. Cluster C had a higher prevalence of any common virus detections compared with Cluster A (Cluster A: 83%, Cluster C: 100%), but this difference was not significant after multiple test correction. Cluster C had a higher prevalence of RV-positive cases (Cluster A: 63%, Cluster C: 91%, *p* < 0.05) and a higher prevalence of cases with RV-C (Cluster A: 37%, Cluster C: 70%, *p* < 0.05) compared with Cluster A.

Cluster C had a higher prevalence of the Multiple (Cluster A: 33%, Cluster C: 50%) and Persistent (Cluster A: 7%, Cluster C: 29%) longitudinal hospital respiratory exacerbation phenotypes compared with Cluster A, and a lower prevalence of the Few phenotype (Cluster A: 53%, Cluster C: 18%), but this difference also lost significance after multiple test correction.

### Differences in recurrence between clusters

Kaplan-Meier survival curves showed differences in the time until the next hospital respiratory exacerbation. The percentage of individuals in each cluster that had re-presented at 1 year was 47% for Cluster A, 57% for Cluster B, and 71% for Cluster C. At 2 years, 50% of Cluster A, 70% of Cluster B, and 82% of Cluster C had re-presented; at 5 years, 53% of Cluster A, 74% of Cluster B, and 88% of Cluster C had re-presented; and at 10 years, 57% of Cluster A, 79% of Cluster B, and 88% of Cluster C had re-presented. The time to 50% re-presentation in each cluster was 1.82 years for Cluster A, 0.75 years for Cluster B and 0.31 years for Cluster C.

Cluster C was found to have a significantly shorter time to recurrence compared to Cluster A (hazard ratio: 2.43, 95% CI: 1.33-4.45, *p* < 0.05) in a univariable Cox proportional hazards model (Figure 1). While there was a trend, the difference between C and B was not significant. Considering the observed differences in the prevalence of common respiratory virus detection and specifically RV detection, we assessed whether the observed differences were the result of clinical factors by repeating the survival analysis including any common respiratory virus-positive respiratory viral swab (hazard ratio: 2.26, 95% CI: 1.22-4.22, *p* < 0.05) and RV-positive respiratory viral swab as additional variables in the Cox model (hazard ratio: 2.13, 95% CI: 1.15-3.92, *p* < 0.05), and found that Cluster C continued to demonstrate a shorter time to recurrence compared with the other clusters. We also assessed the effect of discharge diagnosis, and similarly found that Cluster C had a shorter time to recurrence (hazard ratio: 2.44, 95% CI: 1.33-4.46, *p* < 0.01) even when discharge diagnosis (asthma, wheeze or bronchiolitis) was included as a co-variable (Supplementary Figure 2).

**Figure 1.**
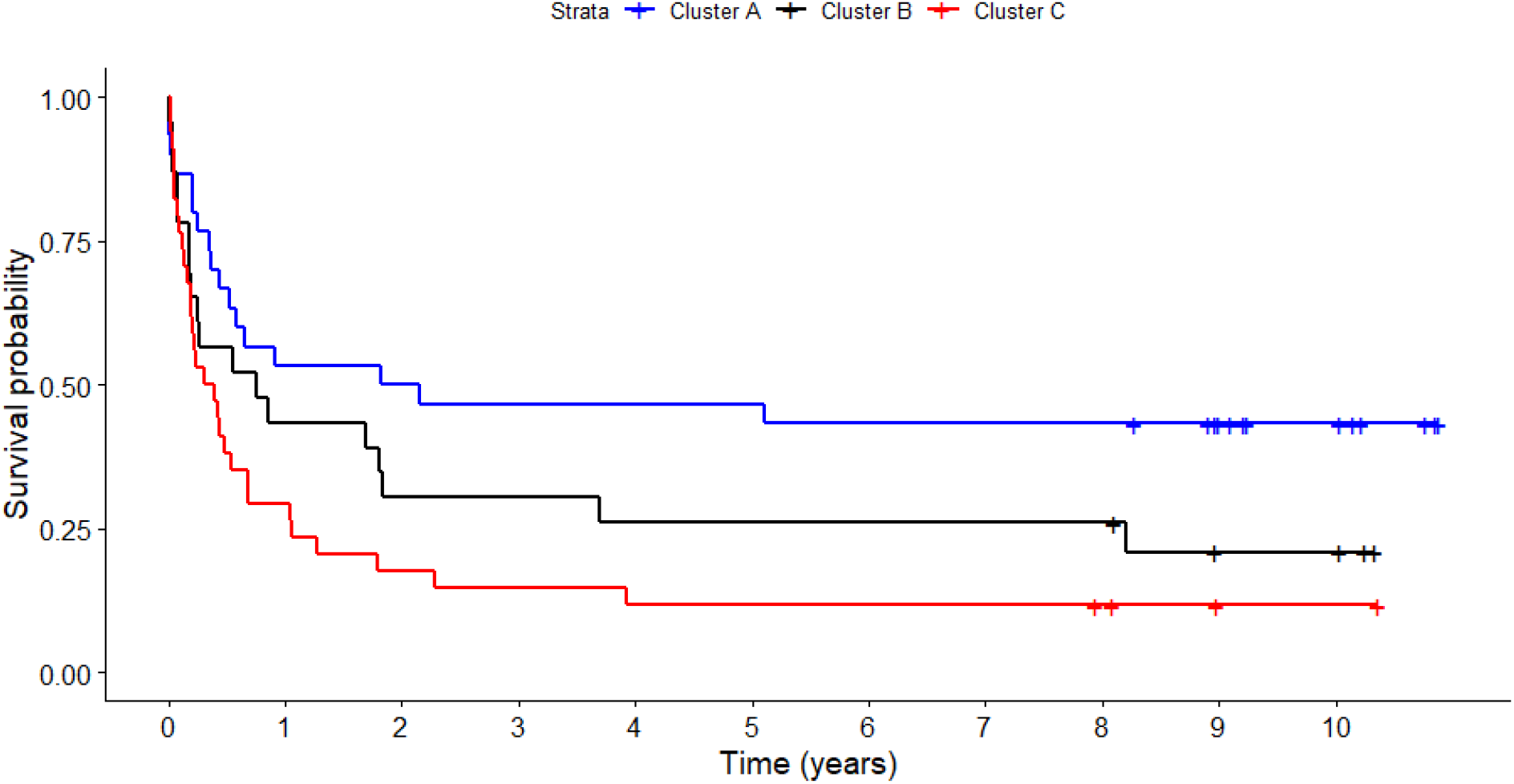
Kaplan-Meier survival curves by Cluster membership. Survival probability indicates the proportion of children yet to experience a subsequent presentation to hospital for a respiratory exacerbation over time measured in years; Clusters A (blue), B (black) and C (red) are the personalised networks method-derived clusters of case children. Crosses indicate censored data.

### Gene module function

To investigate the molecular mechanisms that characterize each cluster, we assessed the relative contributions of the 21 FLAME-derived gene modules to the formation of the 3 clusters. When we considered the top 4 modules based on their mean standardised proportion of network genes in the module, we found that Cluster A was characterised by a higher proportion of Modules 4, 16, 10 and 9 (blue rectangle, Figure 2). Cluster B demonstrated higher proportions of Modules 19, 21, 6 and 7 (black rectangle, Figure 2). Cluster C showed higher portions of Modules 12, 14, 15 and 3 (red rectangle, Figure 2).

**Figure 2.**
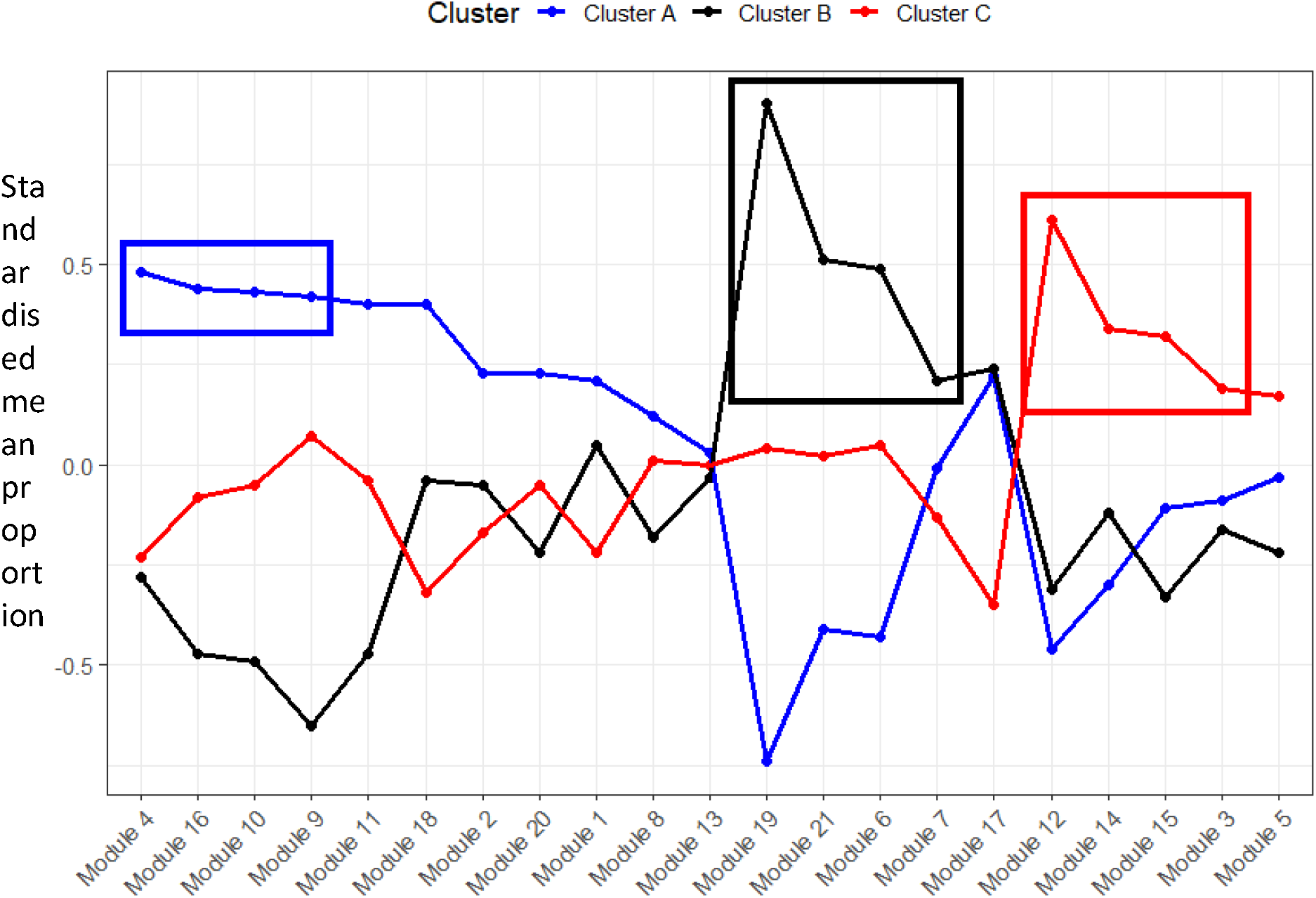
Standardised mean proportion of each FLAME module by cluster. Genes were assigned to modules using FLAME clustering. For each individual, the number of genes in their personal network from each gene module was counted, converted to a proportion of the total network genes and then standardised to account for module size. The mean was then calculated for each cluster. Cluster A = blue, Cluster B = black, Cluster C = red. Boxes highlight the top 4 modules for each cluster.

Table 3 presents the predicted functions from InnateDB and DAVID, and the mean standardised proportions of network genes in each module. Module 4, which had the highest mean proportion in Cluster A, was associated with interferons and anti-viral responses. Modules 16, 10 and 9, also predominant in Cluster A, were associated with innate immunity, immune cell-non-immune cell interactions and Toll-like receptor pathway activation. Modules 19, 21, 6 and 7, which were highest in Cluster B, were associated with G-protein-coupled receptor signalling, glucocorticoid receptor signalling and modulation of histones. Modules 12, 14, 15 and 3, highest in Cluster C, were associated with immunoglobulins, NOTCH1 signalling, TLR signalling, IL12 signalling, NK cells, platelets and apoptosis. Detailed module proportions for each cluster for all modules are available in Supplementary Table 1.

**Table 3:**
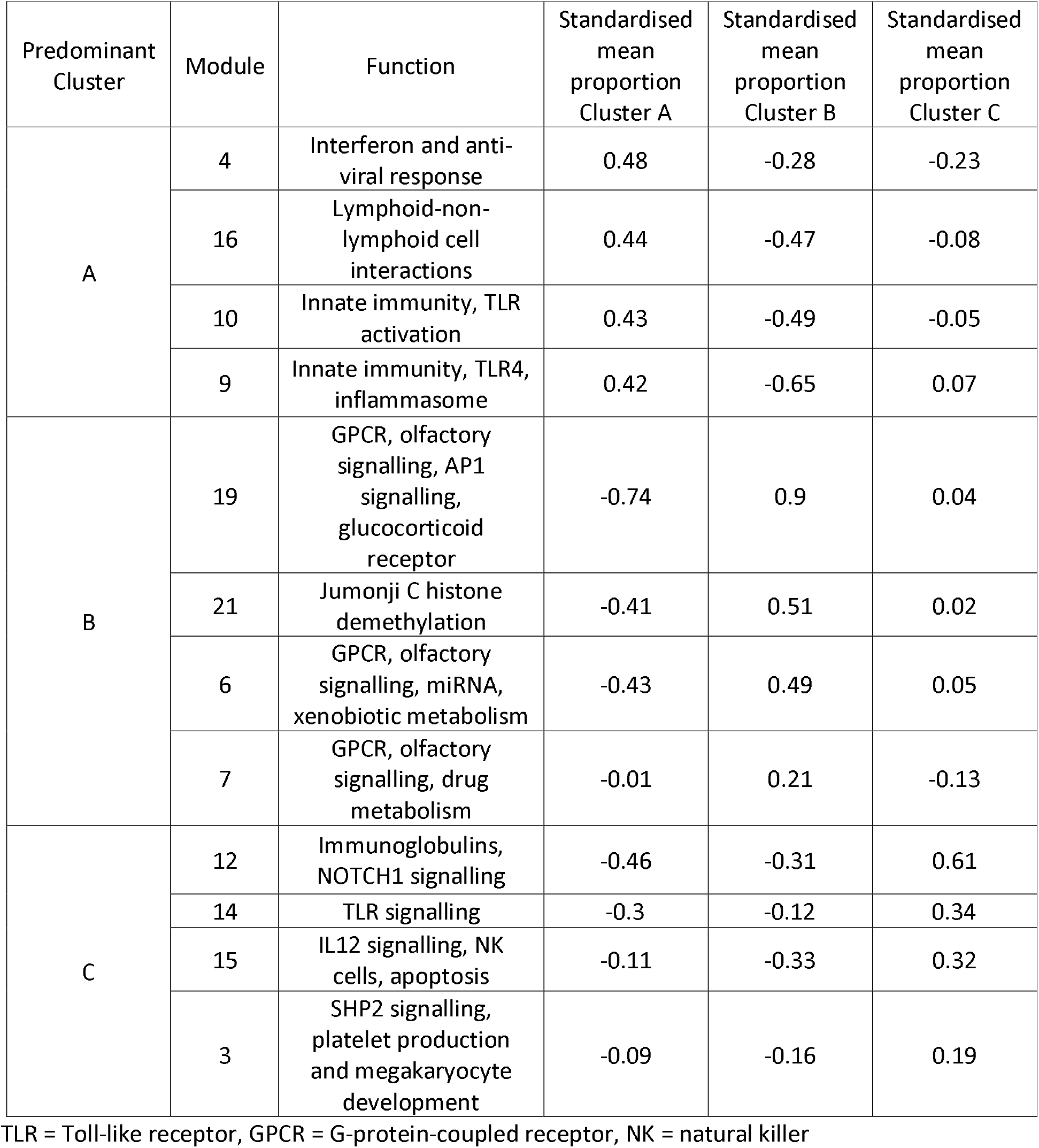
FLAME module functions and proportions in each cluster for top four modules. Genes were assigned to modules using FLAME clustering. For each individual, the number of genes in their personal network from each gene module was counted, converted to a proportion of the total network genes and then standardised to account for module size. The mean was then calculated for each cluster.

The functions of the FLAME-derived gene modules were further investigated using STRING protein interaction networks to visualise prior-knowledge interconnections between module genes. Interaction network diagrams for each of the top four gene modules from Table 3 are shown in Figures 3–5.

**Figure 3.**
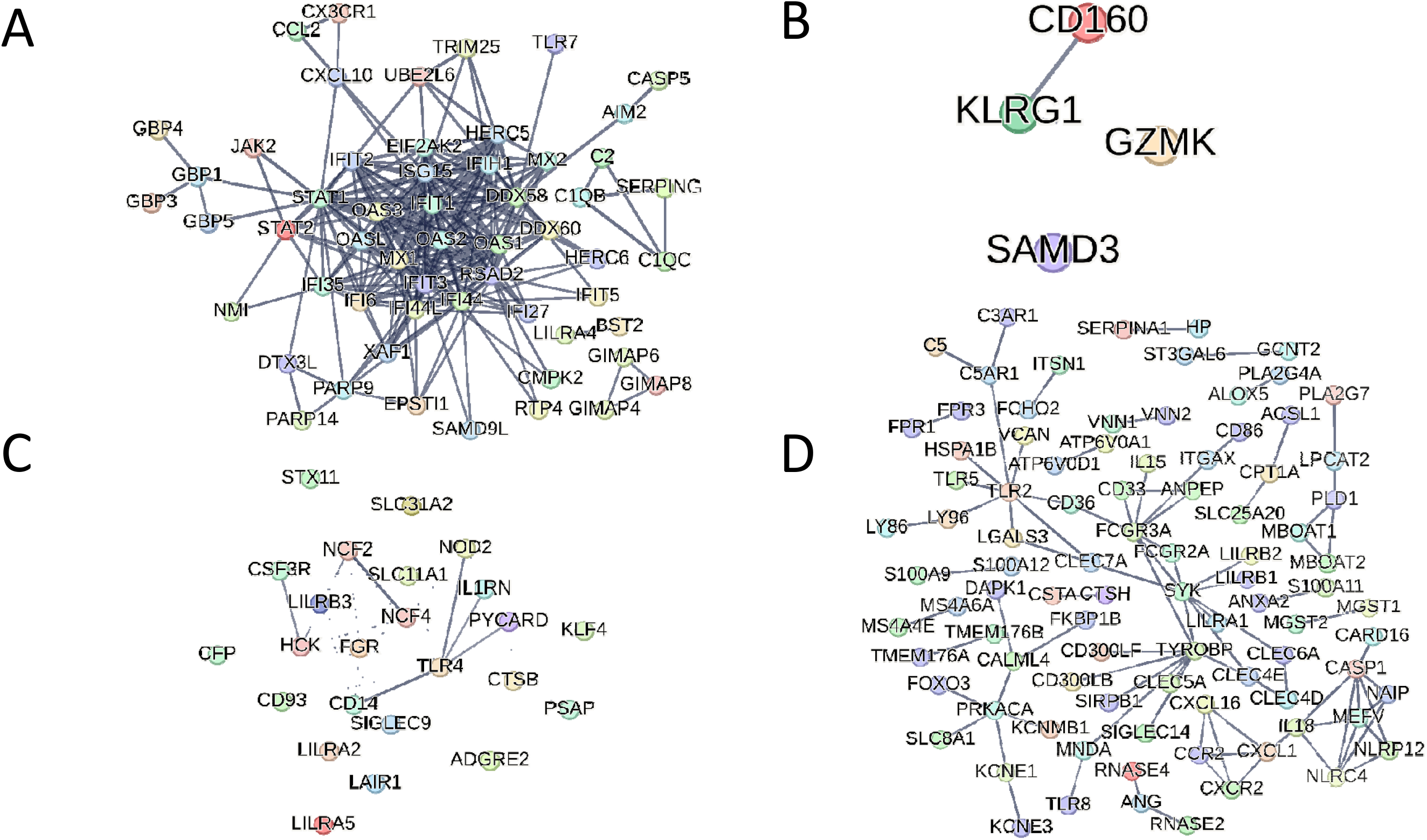
STRING protein interaction networks for the top four FLAME gene modules associated with Cluster A. A) Module 4, B) Module 16, C) Module 10, D) Module 9

**Figure 4.**
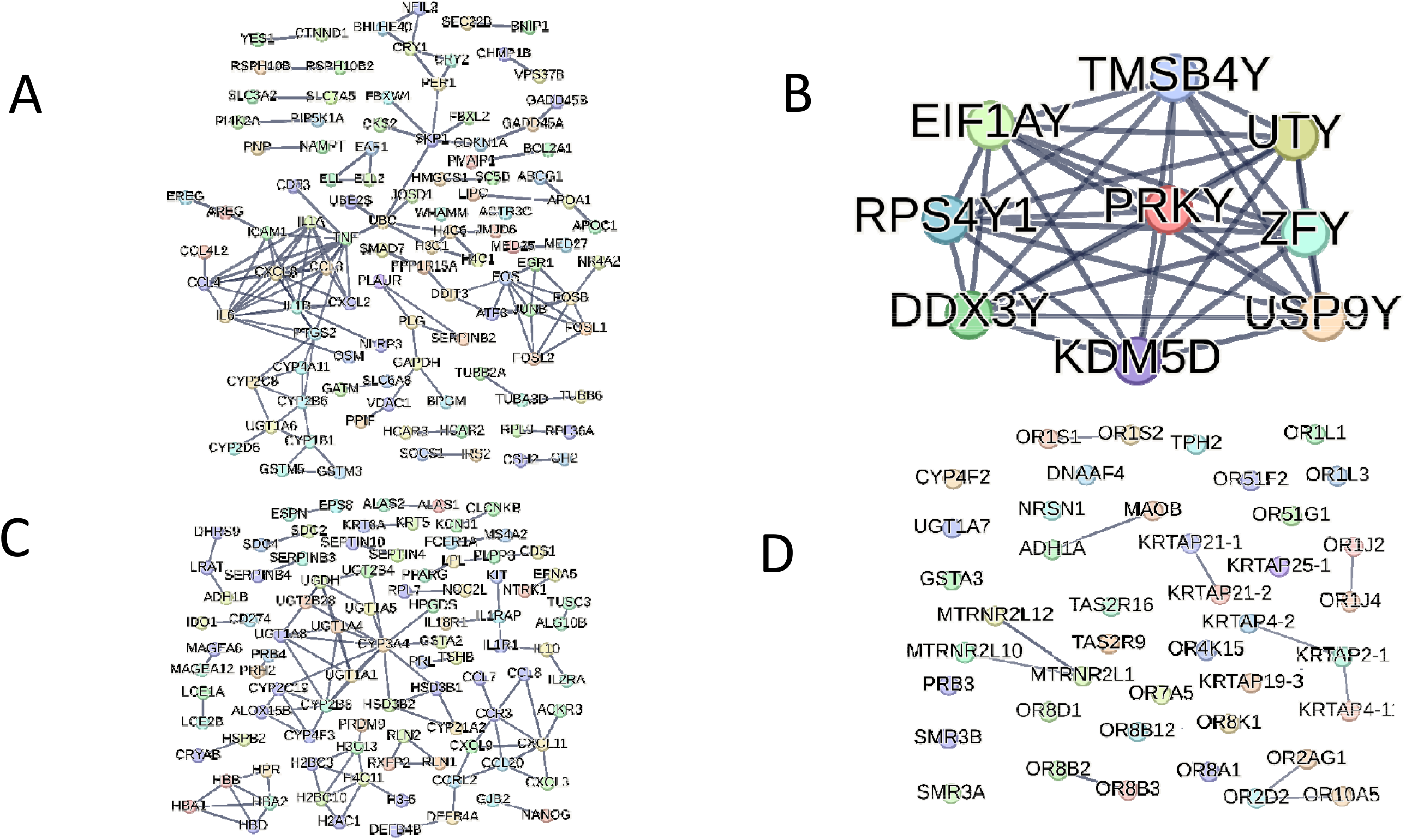
STRING protein interaction networks for the top four FLAME gene modules associated with Cluster B. A) Module 19, B) Module 21, C) Module 6, D) Module 7

**Figure 5.**
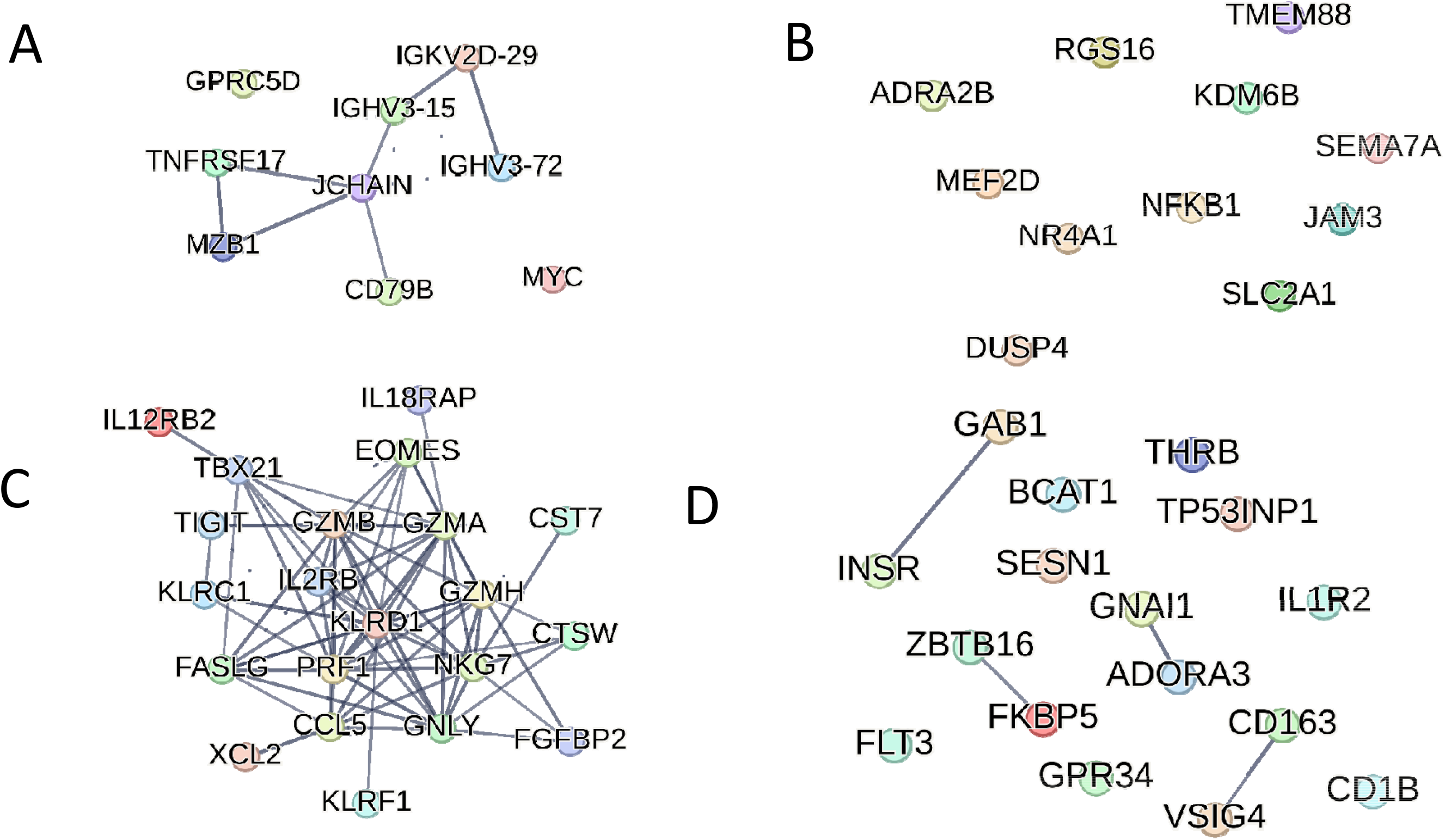
STRING protein interaction networks for the top four FLAME gene modules associated with Cluster C. A) Module 12, B) Module 14, C) Module 15, D) Module 3

## Discussion

In this study, we used a novel method of personal network inference and latent class analysis to categorise children into distinct clusters based on their immune response to an acute lower respiratory illness, particularly acute wheeze and asthma. Our analysis identified three unique clusters, each associated with varying risks of subsequent respiratory-related hospital presentations and the persistent longitudinal respiratory exacerbation phenotype, suggesting these clusters represent functional endotypes. The endotype exhibiting the highest risk of future respiratory hospital exacerbations was characterised by the highest network proportions of gene modules associated with specific innate (TLR, IL12) and adaptive (immunoglobulins) immune pathways, coupled with reduced proportions of interferon-response modules. In contrast, the endotype with the lowest risk of subsequent respiratory hospital presentations had the highest network proportions of gene modules associated with other aspects of innate immunity and interferon responses.

Previous work by our group has identified similar gene expression patterns in the nasal epithelium. Using personal network inference methods analogous to those in this study, we identified endotypes characterised by the balance between interferon- and inflammation-associated genes within each individual’s network, where a low interferon-to-inflammation ratio associated with a reduced time until re-presentation (10). Our current findings suggest that the interferon-low/inflammation-high pattern observed in acutely ill children with lower respiratory illness, often of viral origin, is detectable not only at the site of infection but also systematically in the blood samples.

This is consistent with the previously proposed lung-blood-bone marrow axis model, whereby responses to pathogens at the site of infection in the respiratory tract lead to programming and activation of immune cells in bone marrow that then circulate in blood (25). Flow cytometry analysis of PBMCs collected from 39 children in this cohort showed that plasmacytoid dendritic cells (pDCs) were specifically depleted in children with a persistent hospital respiratory exacerbation phenotype. Moreover, the remaining pDCs exhibited high expression of the high affinity IgE receptor and low expression of interferon-associated genes (14). This finding is, in turn, consistent with a previous small study that suggested that atopy and virus-induced asthma exacerbations may be linked via a mechanism where cross-linking of IgE receptors blocks interferon production in response to viral detection (26). In an animal model of virus/allergen co-exposure-induced asthma, a high-risk rat strain demonstrated high levels of airway inflammation and low expression of interferon-associated genes in lung tissue and bone marrow compared with a low-risk rat strain. Pre-treatment with OM-85, an immunomodulatory agent, ameliorated these effects in high-risk rats by enhancing gene expression in favour of anti-viral genes and reducing eosinophilic inflammation (27). We were not, however, able to assess in this study whether there was concordance between nasal epithelial and blood endotypes as there was little overlap between the subsets assessed. A future study should assess interferon high/low endotypes in both nasal and blood samples from the same participants to address this question.

The present pattern of interferon-high/-low endotypes may help reconcile conflicting findings regarding interferon responses in asthma. Discrepancies in the literature—reporting deficient, elevated, or unchanged interferon levels in asthmatics—could stem from differences in the endotypic composition of study cohorts (28–32). Studies reporting reduced interferon expression may have included a greater proportion of individuals with the interferon-low endotype, whereas those observing elevated or unchanged levels may have had more interferon-high individuals. Our findings linked the interferon-low endotype to persistent hospital exacerbations, suggested that the cohort characteristics, such as frequency of asymptomatic infection, or severity of asthma (stable vs hospitalised), can significantly influence measured interferon levels. An unsupervised cluster analysis of a subset of our cohort showed two distinct groups characterised by high or low expression of interferon-associated genes, with the low-interferon group experiencing a shorter time interval until the next hospital presentation for respiratory illnesses (11). Similarly, in a study of children aged 6-18 years sampled during acute exacerbations, those with lower FEV_1_/FVC ratios—indicative of airflow obstruction—also had reduced expression of interferon-related genes in induced sputum (33). Another study of older children (aged 6-17 years) similarly found that low interferon levels (lowest quartile) in both blood and nasal samples when well were associated with shorter time to the next exacerbation (34). Interestingly, they also found that interferon levels at the time of exacerbation were inversely correlated with FEV_1_ % predicted, indicating potential over-correction in response to a viral infection (34). These previous studies, however, focussed on older children, lacked the ability to characterise individual patterns of gene expression, and, in some cases, did not cluster children based on these expression patterns.

Our results indicate that there are immune endotypes associated with respiratory exacerbations in young children mostly prior to asthma diagnosis. Identifying high-risk immune profiles is crucial for enhancing the management of virus-induced wheeze and asthma exacerbations. This would allow physicians to better inform parents and caregivers regarding future risk management, as well as directing children who will benefit the greatest from more intensive treatment and clinical trials. Clinical trials have explored the efficacy of inhaled interferon-beta in preventing asthma symptoms following viral infections in asthmatic adults. A subgroup analysis indicated that inhaled interferon-beta could prevent loss of asthma control in patients with severe, difficult-to-treat asthma, suggesting potential benefits for individuals who may be interferon-deficient and at risk of persistent severe exacerbations (35).

One interesting observation was that we did not identify gene modules or clusters of children associated with Th2 inflammation, despite over 70% of this cohort exhibiting atopy to at least one allergen. This is notable, as the most well-characterised childhood asthma endotype is driven by atopy and Th2 inflammation (36). We hypothesise that this lack of Th2 signature is due to high baseline prevalence of atopy in this population and the timing of sample collection during a moderate-severe acute exacerbation requiring hospital intervention. While Th2 inflammation plays an important role in community asthma cohorts and in school-aged children (36), it may be less prominent or masked during acute exacerbations, where alternative inflammatory pathways could dominate.

The key strengths of our study is the investigation of immune endotypes in a cohort of young children recruited during acute lower respiratory illness requiring hospital presentation, coupled with comprehensive data on their history of hospital respiratory exacerbations from birth. We used a novel method to examine individual gene expression networks, quantify patterns, and detect endotypes. A limitation of this study is that the analysis was conducted using microarray transcriptomic data rather than sequencing. The study was also conducted at a single site, making replication in cohorts from other locations a priority. The longitudinal exacerbation phenotypes used public hospital record data, and as such omit private hospital visits and only include hospitals within the state. Some children could not be assigned a phenotype, possibly due to living outside of Western Australia or making use of local non-public hospitals.

In summary, we have identified a systemic immune endotype in children with acute lower respiratory illness associated with an increased risk of subsequent hospital respiratory exacerbations. These endotypes present a potential treatable trait of paediatric asthma and wheezing, suggesting treatment options that directly or indirectly enhance interferon production and/or reduce inflammation may be appropriate. Classifying children presenting with wheeze and asthma exacerbations based on their immune profiles may enable tailored management strategies aligned with their future exacerbation risk.

## Data Availability

All data produced in the present study are available online at the Gene Expression Omnibus (GSE298367).

## Funding

This work was supported by the National Health and Medical Research Council [grant number NHMRC APP#1045760], Telethon-Perth Children’s Hospital Research Fund, Wal-yan Respiratory Research Centre, and AstraZeneca.

## Abbreviations

ALRI: acute lower respiratory tract illness
ANOVA: analysis of variance
BIC: Bayesian Information Criterion
CNBC: could not be classified
CSO: Cluster Supporting Object
FEV_1_/FVC: ratio of forced expiratory volume in one second to functional vital capacity
FLAME: Fuzzy clustering by Local Approximation of MEmbership
iCM: iSoft Clinical Manager
LIONESS: Linear Interpolation to Obtain Network Estimates for Single Samples
LPA: Latent Profile Analysis
MAVRIC: Mechanisms of Acute Viral Respiratory Infection in Children
NK: natural killer (cell)
PBMC: peripheral blood mononuclear cell
PCH: Perth Children’s Hospital
pDC: plasmacytoid dendritic cell
PMH: Princess Margaret Hospital
RMA: robust multi-array average RNA – ribonucleic acid
RSV: respiratory syncytial virus
RT-PCR: real-time polymerase chain reaction
RV: rhinovirus
TLR: toll-like receptor

## Notes

### Competing Interest Statement

The authors have declared no competing interest.

### Funding Statement

This study was funded by the National Health and Medical Research Council [grant number NHMRC APP#1045760], Telethon-Perth Children's Hospital Research Fund, Wal-yan Respiratory Research Centre and AstraZeneca.

### Author Declarations

Ethics committee/IRB of Perth Children's Hospital (formerly Princess Margaret Hospital) ethical approval for this work

