## Supplement for "Personal network inference identifies children at risk of recurrent wheezing and asthma"

**Gene module construction**

The FLAME modules were constructed using parameters that resulted in the lowest partition index of 0% cluster overlap and cosine distance as the distance metric. These parameters produced 21 modules, the details and members of which are provided in Supplementary Table 1.

**Personal network construction, module quantification within networks and cluster detection**

We constructed personal gene networks for each subject by selecting the top one percent of edges with the greatest absolute edge weights, as illustrated in Supplementary Figure 1A. Each gene within these networks was annotated according to its FLAME module membership, allowing us to calculate the proportion of network genes corresponding to each FLAME module (Supplementary Figure 1B). These module proportions served as input for LPA, which identified distinct clusters among the participants. The LPA model selection criteria indicated that a variable volume, variable shape, equal orientation (VVE) model with three components - hereafter referred to as Clusters A, B and C - optimised the BIC (Supplementary Figure 1C).

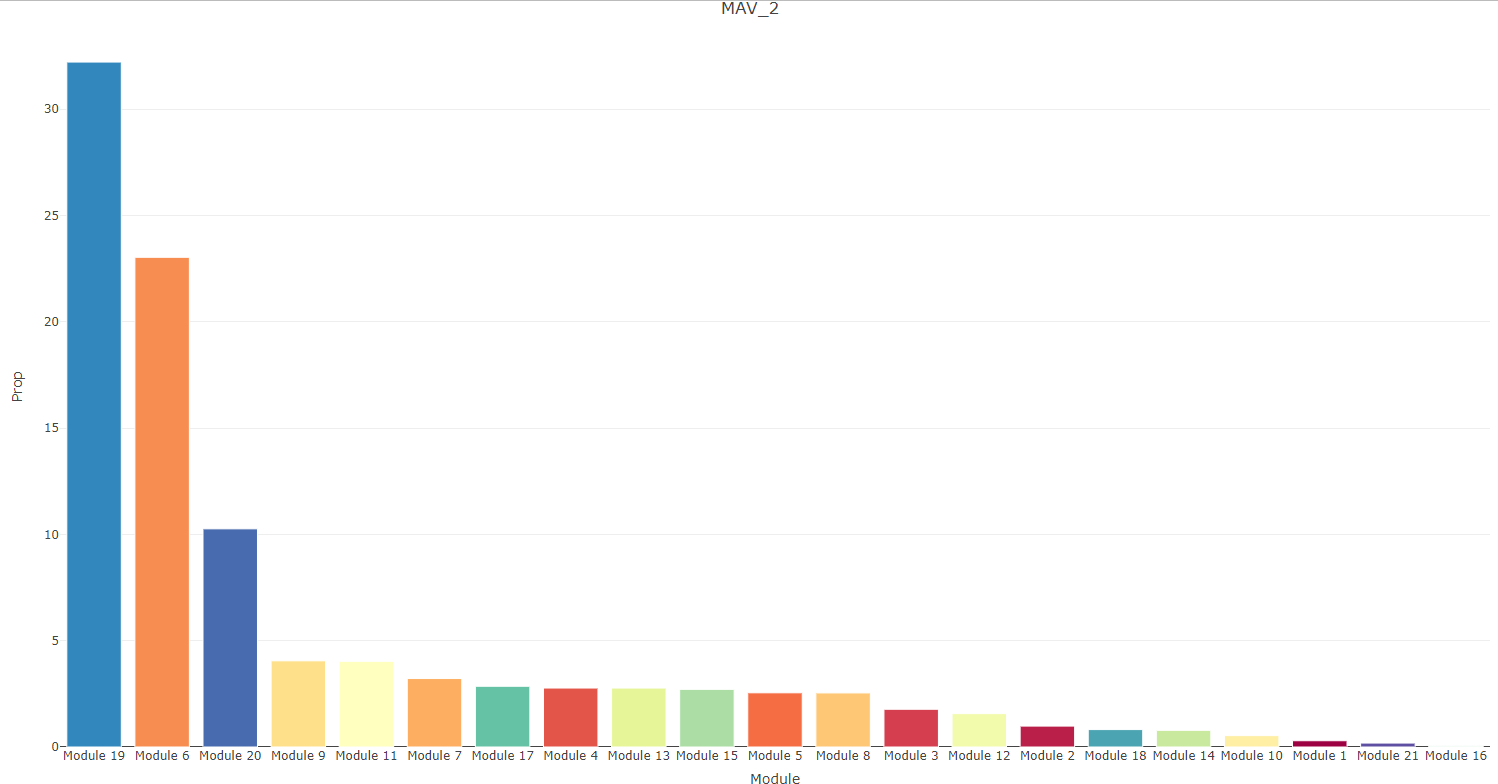

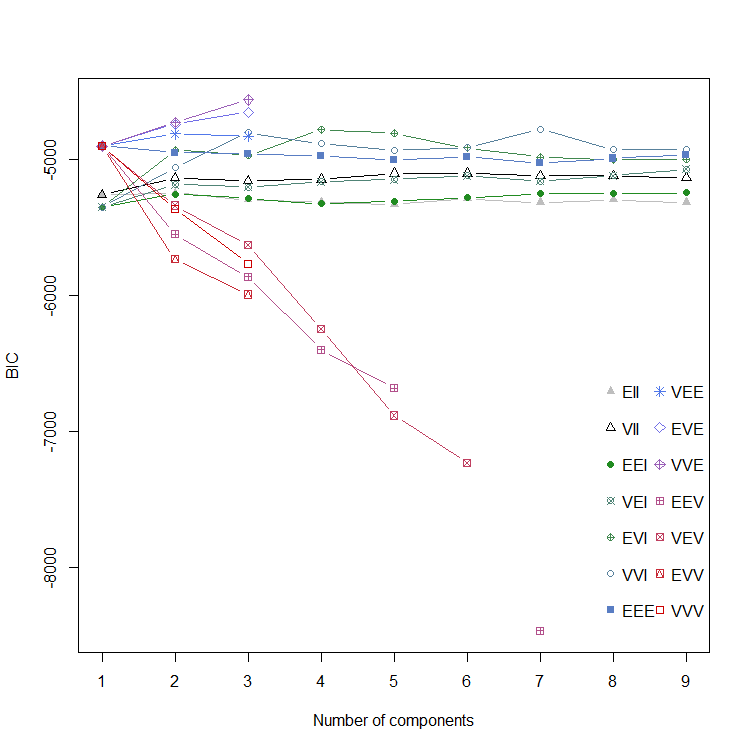

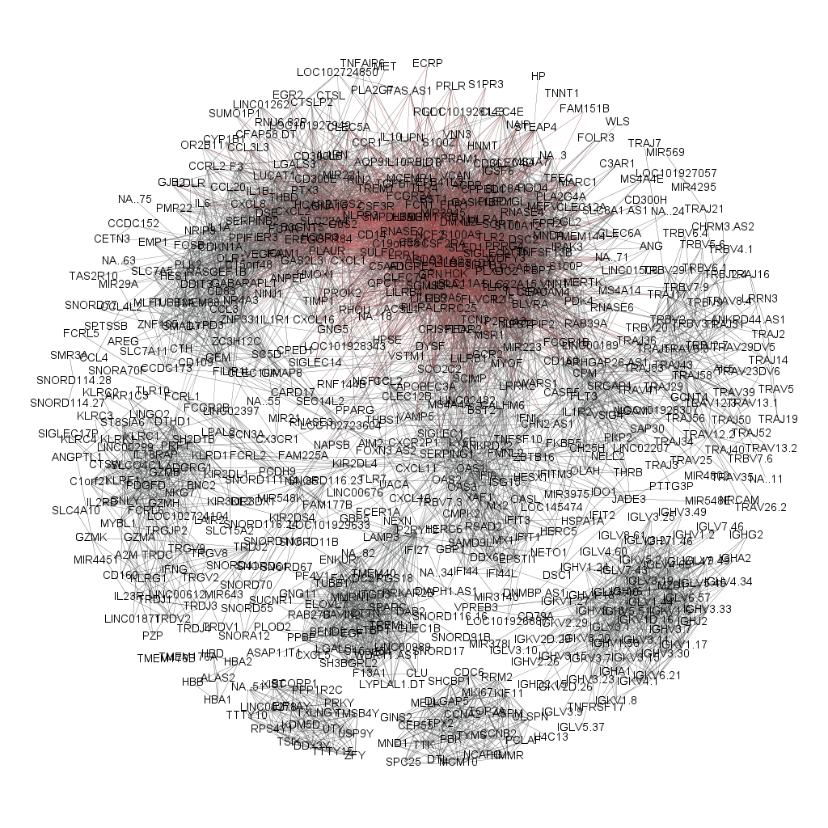

A

B

C

*

**Supplementary Figure 1:** Personalised network methodology. A) Example of a one percent gene expression network from Gephi, where red indicates the genes with the most connections to other genes in the network, the blue circle indicates an interferon-associated gene module, the green circle indicates an inflammation-associated gene module B) example of proportions (%) of network genes in each FLAME module, calculated by summing the number of one percent gene network members from each FLAME module, C) Latent Profile Analysis model selection, number of components (clusters) versus Bayesian Information Criterion (BIC), separated by model type. The best model is that which maximises the BIC. V = variable, E = equal, I = undefined. The best model is indicated by *

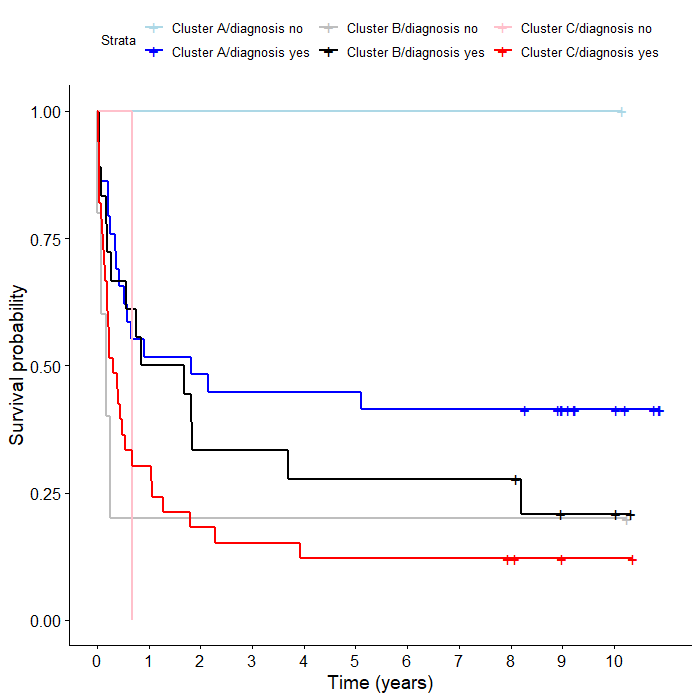

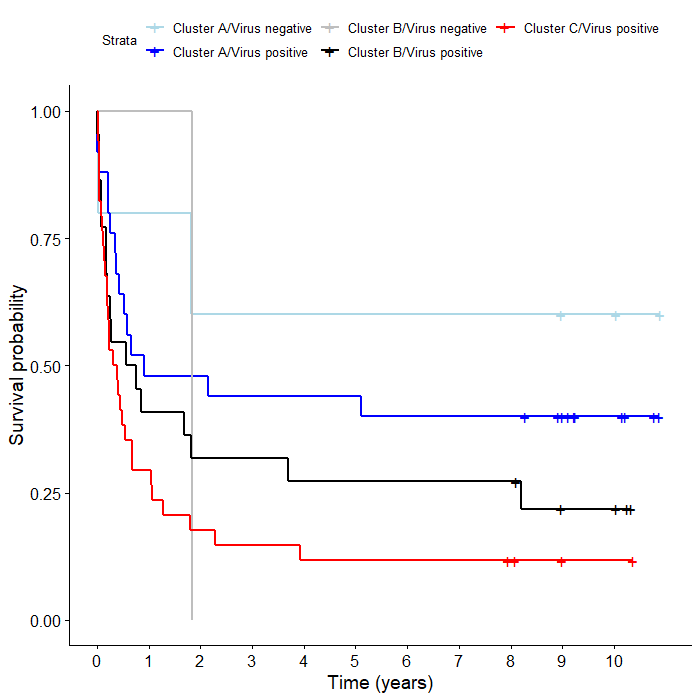

A

C

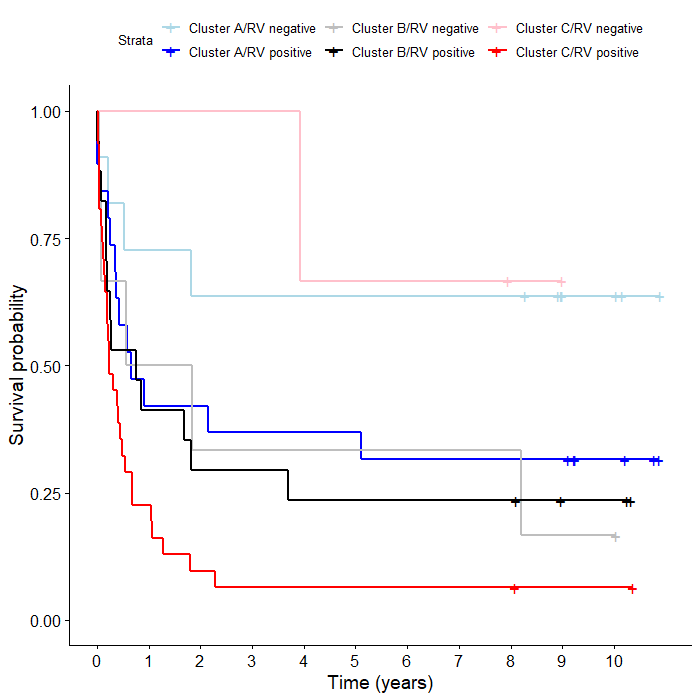

B

**Supplementary Figure 2:** Kaplan-Meier survival curves separated by Cluster membership and virus detection or discharge diagnosis. Survival probability indicates the proportion of children yet to experience a subsequent presentation to hospital for a respiratory exacerbation over time measured in years; Clusters A, B and C are the personalised networks method-derived clusters of case children. A) any common respiratory virus-positivity, B) RV-positivity, C) discharge diagnosis of acute asthma, wheeze or bronchiolitis. Crosses indicate censored data.

**Supplementary Table 1:** FLAME module functions and standardised mean proportions in each cluster

| Module | Function | Standardised mean proportion Cluster A | Standardised mean proportion Cluster B | Standardised mean proportion Cluster C | Predominant Cluster |
| --- | --- | --- | --- | --- | --- |
| 1 | Platelet activation and blood coagulation | 0.21 | 0.05 | -0.22 | A |
| 2 | Platelets, vascular smooth muscle contraction, endocytosis | 0.23 | -0.05 | -0.17 | A |
| 3 | SHP2 signalling, platelet production and megakaryocyte development | -0.09 | -0.16 | 0.19 | C |
| 4 | Interferon and anti-viral response | 0.48 | -0.28 | -0.23 | A |
| 5 | Immunoglobulins, NK cells | -0.03 | -0.22 | 0.17 | C |
| 6 | GPCR, olfactory signalling, miRNA, xenobiotic metabolism | -0.43 | 0.49 | 0.05 | B |
| 7 | GPCR, olfactory signalling, drug metabolism | -0.01 | 0.21 | -0.13 | B |
| 8 | Cell cycle/mitosis, histones | 0.12 | -0.18 | 0.01 | A |
| 9 | Innate immunity, TLR4, inflammasome | 0.42 | -0.65 | 0.07 | A |
| 10 | Innate immunity, TLR activation | 0.43 | -0.49 | -0.05 | A |
| 11 | Lysosome, inflammation | 0.4 | -0.47 | -0.04 | A |
| 12 | Immunoglobulins, NOTCH1 signalling | -0.46 | -0.31 | 0.61 | C |
| 13 | B cell signalling | 0.03 | -0.03 | 0.00 | A |
| 14 | TLR signalling | -0.3 | -0.12 | 0.34 | C |
| 15 | IL12 signalling, NK cells, apoptosis | -0.11 | -0.33 | 0.32 | C |
| 16 | Lymphoid-non-lymphoid cell interactions | 0.44 | -0.47 | -0.08 | A |
| 17 | Histones, nucleosome | 0.22 | 0.24 | -0.35 | B |
| 18 | Nucleolus, RNA processing | 0.4 | -0.04 | -0.32 | A |
| 19 | GPCR, olfactory signalling, AP1 signalling, glucocorticoid receptor | -0.74 | 0.9 | 0.04 | B |
| 20 | T cells, immunoglobulins | 0.23 | -0.22 | -0.05 | A |
| 21 | Jumonji C histone demethylation | -0.41 | 0.51 | 0.02 | B |

GPCR = G-protein-coupled receptor, NK = natural killer, TLR = Toll-like receptor
